# Clinical Validity of Autosomal Dominant *ALPK3* Loss-of-function Variants as a Cause of Hypertrophic Cardiomyopathy

**DOI:** 10.1101/2025.03.27.25324722

**Authors:** Sophie Hespe, Emma S. Singer, Chloe Reuter, Brittney Murray, Elizabeth Jordan, Jessica Chowns, Stacey Peters, Megan Mayers, Belinda Gray, Ray E. Hershberger, Anjali Owens, Christopher Semsarian, Amber Waddell, Babken Asatryan, Emma Owens, Courtney Thaxton, Mhy-Lanie Adduru, Kailyn Anderson, Emily E. Brown, Lily Hoffman-Andrews, Fergus Stafford, Richard D. Bagnall, Lucas Bronicki, Bert Callewaert, C. Anwar A. Chahal, Cynthia A. James, Olga Jarinova, Andrew P. Landstrom, Elizabeth M. McNally, Laura Muiño-Mosquera, Victoria Parikh, Roddy Walsh, Bess Wayburn, James S. Ware, Benjamin L. Parker, Enzo R. Porrello, David A. Elliott, James W. McNamara, Jodie Ingles

## Abstract

*ALPK3* encodes the protein α-kinase 3, an essential cardiac-enriched atypical a-kinase that inserts in the nuclear envelope and the sarcomere M-band of cardiac myocytes, functioning to aid in myosin-mediated force buffering and sarcomere proteostasis. Previously, bi-allelic loss-of-function *ALPK3* variants have been reported causative in a severe paediatric phenotype including hypertrophic (HCM) and dilated cardiomyopathy (DCM). Very few heterozygous carries in these cases express any cardiac phenotype. However, recently studies have reported heterozygous loss-of-function *ALPK3* variants causative of HCM. In this research letter we present a patient series of 29 cardiac patients (26 probands) with heterozygous putative loss-of-function *ALPK3* variants without another causative variant in other definitive HCM genes. As well as, a ClinGen gene curation assessing the clinical validity of the gene-disease association of *ALPK3* for autosomal dominant HCM, which was evaluated to be strong. With reduced penetrance compared to other HCM genes and issues with historic genetic reports using a superseded transcript care needs to be taken when interpreting these variants to account for these nuances.

## Introduction

*ALPK3* encodes the protein α-kinase 3, an essential cardiac-enriched atypical a-kinase that inserts in the nuclear envelope and the sarcomere M-band of cardiac myocytes, functioning to aid in myosin-mediated force buffering and sarcomere proteostasis.^1^ In 2016, bi-allelic variants in *ALPK3* were reported as a cause of paediatric cardiomyopathy including hypertrophic (HCM) and dilated cardiomyopathies (DCM), with extracardiac features such as musculoskeletal abnormalities, facial dysmorphism and cleft palate.^2^ At the time, very few heterozygous carrier family members expressed any cardiac phenotype. Recently, heterozygous putative loss-of-function (LOF) *ALPK3* variants have been reported as a cause of HCM, potentially accounting for 1-2% of adult cases based on burden testing.^3^ In addition, a locus near *ALPK3* has been associated with both HCM and DCM in genome-wide association studies, indicating that common variants might contribute to cardiomyopathy risk.

*ALPK3* is located at ch15q25.3 and has 14 exons encoding 1,705 amino acids on the most recent transcript (NM_020778.5). The previous transcript (NM_020778.4) encoded 1,907 amino acids, which included an additional 201 amino acids preceding the start of exon 1, and one amino acid at the end of exon 14 (Figure 1C). Curation of bi-allelic *ALPK3* variants previously showed definitive evidence for autosomal recessive infantile-onset cardiomyopathy. The Clinical Genome Resource (ClinGen) Hereditary Cardiovascular Disorders (HCVD) Gene Curation Expert Panel determined the autosomal dominant (AD) *ALPK3* disease entity should be split from childhood-onset autosomal recessive *ALPK3* cardiomyopathy and applied the ClinGen framework for evaluating gene-disease clinical validity.^4^ Here we report a summary of new and existing evidence contributing to the ClinGen HCVD Gene Curation Expert Panel classification of AD HCM, with attention to the real-world challenges noting the large proportion of patients this may impact. During the ClinGen gene curation process, anecdotal inconsistencies and challenges in considering AD *ALPK3* variants as a cause of HCM were discussed, and we have highlighted these by compiling an international patient series to illustrate the nuance of our curation.

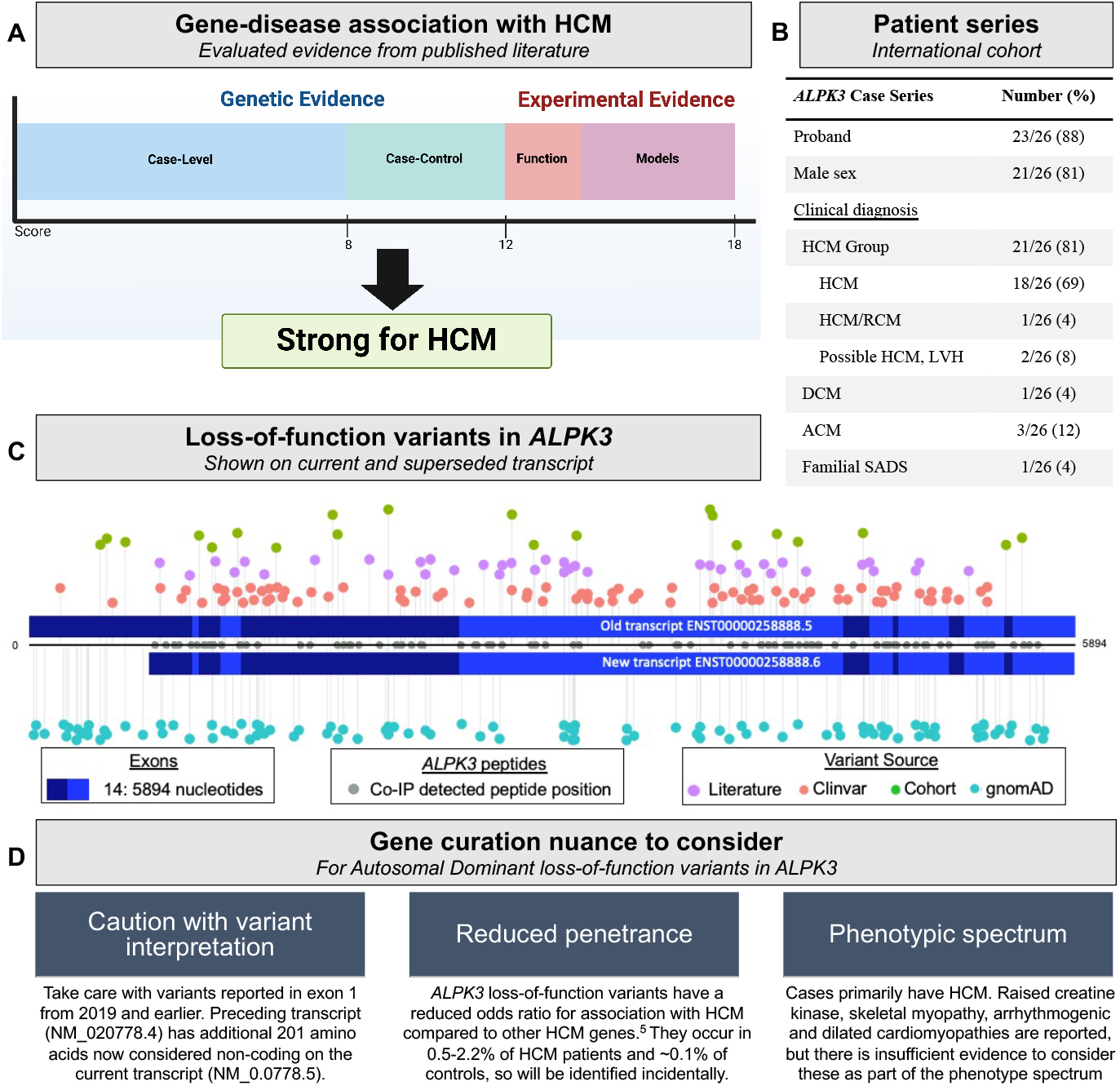
CENTRAL ILLUSTRATION: (A) Summary of ClinGen autosomal dominant *ALPK3* loss-of-function (LOF) curation for hypertrophic cardiomyopathy (HCM); (B) Characteristics of patients with heterozygous *ALPK3* LOF variants from the international patient series; (C) Gene topology of *ALPK3* LOF variants; (D) Autosomal dominant *ALPK3* gene curaction key points. Gene topology of *ALPK3* showing exons, defined for the old transcript (ENST00000258888.5; NM_020778.4), and the new transcript (ENST00000258888.6; NM_020778.5). Coding regions of both transcripts were plotted and aligned by nucleotide position, with ENST00000258888.5 starting at 0 and ending at 5894, and ENST00000258888.6 starting at position 677 and ending at 5891. Co-immunoprecipitation studies in human pluripotent stem cell derived-cardiomyocytes showed ALPK3 peptides matching only the revised transcript, plotted in grey. *ALPK3* LOF variants in the case series (green); ClinVar (red; November 2022), literature (purple) and the Genome Aggregation Database (light blue; gnomAD v2.1; November 2022) were extracted and plotted along with our cohort variants. Abbreviations: Co-IP, Co-immunoprecipitation; HCM, hypertrophic cardiomyopathy; DCM, dilated cardiomyopathy; ACM, arrhythmogenic cardiomyopathy; RCM, restrictive cardiomyopathy; LVH, left ventricular hypertrophy; SADS, sudden arrhythmic death syndrome.

### *ALPK3* patient series

Twenty-nine patients (26 probands) with heterozygous putative loss-of-function (LOF) *ALPK3* variants and without another causative variant in other definitive HCM genes from seven institutions in Australia, USA, and the UK were included. Human research ethical approval was granted at each site according to local regulations. LOF variants included frameshift, nonsense, and splice site, predicted to result in nonsense mediated decay. Clinical diagnosis and family history data were collated. Three patients had variants reported on the superseded transcript (now non-coding) and were excluded. The remaining 26 patients (23 probands) had 18 unique variants and clinical diagnoses including; HCM group (including HCM/RCM and possible/borderline diagnoses; n=21; 80%), DCM (1; 4%), arrhythmogenic cardiomyopathy (3; 12%), and sudden arrhythmic death (1; 4%) (Figure 1B).

### *ALPK3* gene curation

Gene curation was performed by the ClinGen HCVD Gene Curation Expert Panel as part of the re-appraisal of HCM gene validity.^4^ The full summary and citations are available online (https://search.clinicalgenome.org/CCID:008323). *ALPK3* was first reported in relation to autosomal dominant HCM in 2018. At least 38 variants (nonsense, frameshift, and splice site) have been reported in probands in >5 publications. Variants segregated in at least 3 families (summed LOD score of 1.8). Five case-control studies reported the frequency of *ALPK3* LOF variants in HCM cohorts (0.5-2.2%), being significantly greater than control cohorts (0.07-0.15%), including European, Chinese and Korean ancestry populations. More evidence was available in the literature, however the maximum score for genetic evidence (12 points) was reached. Experimental evidence included biochemical function, expression, and models, reaching the maximum score of 6 with the inclusion of a recent knock-in mouse model which demonstrated a hypertrophic response following chronic adrenergic challenge, and partial rescue of the cellular phenotype by Mavacamten. Overall, *ALPK3* was classified as strong (score of 18) for autosomal dominant HCM (Figure 1A).

## CLINICAL IMPLEMENTATION

Heterozygous LOF *ALPK3* variants are enriched in cases compared to controls, and may contribute to approximately 0.5-2.2% of adult-onset HCM. Reported phenotype includes more frequent apical or concentric patterns of hypertrophy, myocardial fibrosis and shorter PR interval, compared to sarcomere-positive HCM patients.^3^ The same study showed 20% (7/35) of patients have raised serum creatine kinase, and 1 patient had clinically apparent skeletal involvement. Reduced penetrance of AD LOF *ALPK3* variants has been suggested relative to *MYBPC3* variants and there should be a high degree of caution when interpreting these as secondary genetic findings.^5^ Patients with other cardiac phenotypes including DCM and arrhythmogenic cardiomyopathy have been described, though there is currently insufficient evidence to consider these as part of the phenotype spectrum, and may relate to reduced penetrance of the variants (i.e., secondary findings) (Figure 1D).

As highlighted in our patient series, when reviewing historic genetic reports (prior to September 2019), care should be taken due to a superseded transcript, meaning some exon 1 variants are now considered non-coding. Indeed, mapping the peptides from endogenous *ALPK3* enriched by co-immunoprecipitation in human pluripotent stem cell-derived cardiomyocytes matched the revised transcript further supporting its importance in cardiomyocytes (PXD035734).

## Data Availability

All data produced in the present study are available upon reasonable request to the authors

## ACKNOWLEDGEMENTS/FUNDING

JI is the recipient of a National Heart Foundation of Australia Future Leader Fellowship (#106732). CS is the recipient of a National Health and Medical Research Council (NHMRC) Investigator Grant (#2016822) and a New South Wales Health Cardiovascular Disease Clinician Scientist Grant. RDB is the recipient of a New South Wales Cardiovascular Disease Senior Scientist Grant. JSW is supported by Medical Research Council (UK), Sir Jules Thorn Charitable Trust [JTA1], British Heart Foundation [RE/18/4/34215], and the NIHR Imperial College B--iomedical Research Centre. JWM is the recipient of a National Heart Foundation of Australia Future Leader Fellowship (#107192) and Medical Research Future Fund Stem Cell Mission (MRF2024440).

## Notes

### Competing Interest Statement

EMM consults for Amgen, Cytokinetics, PepGen, Tenaya Therapeutics and is a founder of Ikaika Therapeutics and EnCarda. JW has consulted for MyoKardia, Inc., Pfizer, Foresite Labs, Health Lumen, and Tenaya Therapeutics, and receives research support from Bristol Myers-Squibb. JI receives research grant support from Bristol Myers Squibb unrelated to this work. CAJ receives research funding from Lexeo Therapeutics, Arvada Therapeutics, EicOsis, and StrideBio Inc and has consulted for Pfizer and Lexeo Therapeutics. E.R.P. are cofounders, scientific advisors and hold equity in Dynomics, a biotechnology company focused on the development of heart failure therapeutics. All other authors declare no competing interests.

### Author Declarations

Each participating site has received ethics approval in accordance with local policies, as per the following: ethical approval requiring informed consent was obtained from Childrens Hospital of Philadelphia USA, Yale Medical USA, Royal Brompton Hospital United Kingdom, Melbourne Health Human Research Australia, and Sydney Local Health District Royal Prince Alfred Hospital Australia ethics committees. Waiver of consent was granted by Stanford School of Medicine USA, Pennsylvania University Medical Center USA, Johns Hopkins University USA, and Sydney Local Health District Royal Prince Alfred Hospital Australia ethics committees.

